# Ad hoc laboratory-based surveillance of SARS-CoV-2 by real-time RT-PCR using minipools of RNA prepared from routine respiratory samples

**DOI:** 10.1101/2020.03.30.20043513

**Authors:** Anna M. Eis-Hübinger, Mario Hönemann, Jürgen J. Wenzel, Annemarie Berger, Marek Widra, Barbara Schmidt, Souhaib Aldabbagh, Benjamin Marx, Hendrik Streeck, Sandra Ciesek, Uwe. G. Liebert, Daniela Huzly, Hartmut Hengel, Marcus Panning

## Abstract

We report a laboratory-based surveillance for SARS-CoV-2 using minipools of respiratory samples submitted for routine diagnostics. We tested a total of 70 minipools resembling 700 samples shortly before the upsurge of cases in Germany. We identified one SARS-CoV-2 positive patient. Our approach proved its concept, is easily adaptable and resource-saving.

## Text

As of 11 March 2020, WHO declared COVID-19 a pandemic (1). Early case detection is crucial to contain the pandemic and symptom-based case definitions have been set up in many countries worldwide. However, there is evidence that transmission chains can be initiated by asymptomatic cases or only mildly diseased COVID-19 patients (2). These cases will be missed by currently recommended symptom-based case definitions and may lead to unrecognized local spread, which has been seen in Italy, Iran and more recently in parts of the US. One of the biggest challenges and unresolved issues for public health is the rapid identification of SARS-CoV-2 transmission chains within the general population and ultimately in hospitals.

Here we propose an *ad hoc* laboratory-based surveillance approach for SARS-CoV-2 which might help to identify unrecognized spread in an efficient, resource-saving and cost effective manner. It is based upon minipool (MP) testing of nucleic acid preparations of respiratory samples submitted to laboratories for routine diagnostics.

## The study

The workflow comprises individual nucleic acid (NA) extraction of respiratory samples, pooling of extracted NA samples in batches of 10 and SARS-CoV-2 specific real-time RT-PCR. In a first step, we analyzed the impact of minipool (MP) testing in batches of 10 samples per pool. Nucleic acid was extracted from 200 µl respiratory specimen (pharyngeal swabs in viral transport medium, sputum, broncho-alveolar lavage fluid) using the MinElute Virus kit (Qiagen, Hilden, Germany) on the QIAcube system as recommended. Elution was done in a volume of 100 µl. For setting up MP, 5 µl of each individual NA preparation was combined in pools of 10 (dilution factor of 10). We retrieved 40 left-over NA preparations of respiratory samples representing a variety of non-SARS-CoV-2 viruses from our local biobank in Freiburg and set up MP. We tested four MP using the same RT-PCR as for individual patient testing as described (3). We were able to detect all viral pathogens which tested positive in individual RT-PCR (Table 1). To exclude possible unspecific reactions of the MP procedure these MP were also tested using the SARS-CoV-2 specific real-time RT-PCR as described below and no unspecific reactions were observed.

**Table 1:** Detection of respiratory viruses in samples using individual RT-PCR and in four minipools of 10 individual samples (A1 – A4), Freiburg, Germany, December 2019.

| Patient sample | Pathogen | Ct-value<br>(Individual patient analysis) | Minipool | Pathogen | Ct-value<br>(Minipool analysis) |
| --- | --- | --- | --- | --- | --- |
| 1 | Influenza B virus | 29 | A1 | Influenza B virus | 25 |
| 2 | negative |  |  | negative |  |
| 3 | negative |  |  | negative |  |
| 4 | negative |  |  | negative |  |
| 5 | negative |  |  | negative |  |
| 6 | negative |  |  | negative |  |
| 7 | negative |  |  | negative |  |
| 8 | negative |  |  | negative |  |
| 9 | negative |  |  | negative |  |
| 10 | negative |  |  | negative |  |
| 11 | negative |  | A2 | negative |  |
| 12 | RSV | 25 |  | RSV | 29 |
| 13 | negative |  |  | negative |  |
| 14 | negative |  |  | negative |  |
| 15 | Influenza A virus | 33 |  | Influenza A virus | 34 |
| 16 | negative |  |  | negative |  |
| 17 | negative |  |  | negative |  |
| 18 | negative |  |  | negative |  |
| 19 | negative |  |  | negative |  |
| 20 | negative |  |  | negative |  |
| 21 | negative |  | A3 | negative |  |
| 22 | Rhinovirus, HMPV | 24, 25 |  | Rhinovirus, HMPV | 31, 30 |
| 23 | negative |  |  | negative |  |
| 24 | Adenovirus | 25 |  | Adenovirus | 29 |
| 25 | negative |  |  | negative |  |
| 26 | negative |  |  | negative |  |
| 27 | negative |  |  | negative |  |
| 28 | RSV | 32 |  | RSV | 35 |
| 29 | Negative |  |  | negative |  |
| 30 | negative |  |  | negative |  |
| 31 | negative |  | A4 | negative |  |
| 32 | RSV | 34 |  | RSV | >35 |
| 33 | Influenza A virus | 37 |  | Influenza A virus | 33 |
| 34 | negative |  |  | negative |  |
| 35 | Influenza A virus | 32 |  | Influenza A virus | 29 |
| 36 | negative |  |  | negative |  |
| 37 | negative |  |  | negative |  |
| 38 | negative |  |  | negative |  |
| 39 | negative |  |  | negative |  |
| 40 | HMPV | 32 |  | HMPV | 34 |

To determine the analytical sensitivity of the MP approach, we used *in vitro*-transcribed RNA standards for the E gene obtained by the European virus archive global (EVAg), https://www.european-virus-archive.com, and the SARS-CoV-2 E gene RT-PCR assay as described (4). RT-PCR was done on an ABI 7500 instrument (Applied Biosystems, Weiterstadt, Germany). We spiked different *in vitro*-transcribed RNA concentrations in stored NA preparations of respiratory samples from 2019 and established MP. Replicate testing was done to determine the limit of detection (LOD) as described (4). The LOD for the MP approach was 48 copies per reaction (95% confidence interval: 33 – 184) (Figure 1). We used NA preparations from three actual SARS-CoV-2 cases in Freiburg (containing 4×10^4^ copies/ml; 3.2×10^7^ copies/ml; 1.6×10^7^ copies/ml, respectively) and set up three MP each containing one SARS-CoV-2 positive NA preparation and retested these samples. Except for the MP containing the low concentrated sample both other MP tested positive.

**Figure 1:**
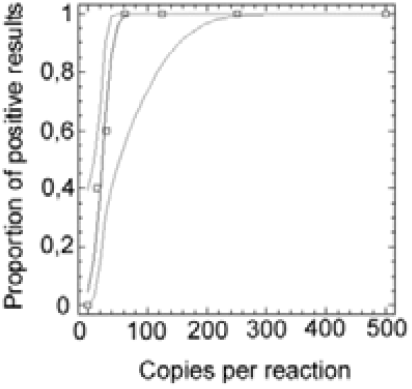
Probit analysis of SARS-CoV-2 RNA detection rate (y axes) in relation to viral RNA concentration at different copy numbers per reaction (x axes).

Finally, we prospectively analyzed 42 MP comprising 420 samples using the SARS-CoV-2 E gene assay. We used all available NA samples which had been sent for routine diagnostics to the Institute of Virology in Freiburg excluding samples with a specific request for SARS-CoV-2 diagnostics from 17.02.2020 to 10.03.2020 (Figure 2). One out of 42 MP tested positive. The MP was resolved and individual testing confirmed SARS-CoV-2 infection in one individual patient.

**Figure 2:**
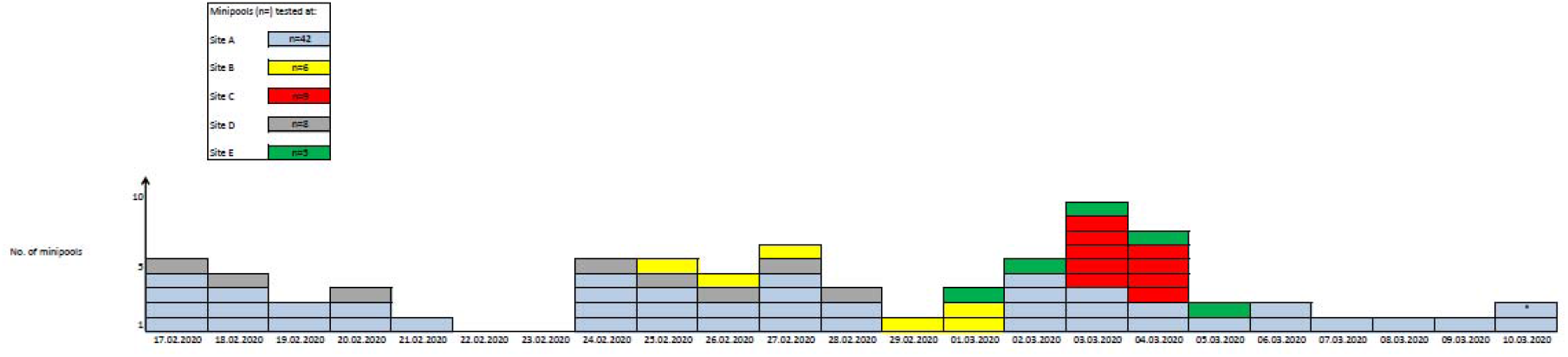
Number of minipools tested by date at five sites in Germany, February-March. *: First SARS-CoV-2 RNA positive minipool detected

We distributed the workflow within an informal network of 5 German laboratories (Table 2). All sites are tertiary care centers with a total of 1.600 (site A), 1.300 (site B), 1.400 (site C), 840 beds (site D), and 1.500 (site E), respectively. Invited laboratories rapidly adopted the MP screening strategy and a total of 70 MP were tested from 17.02.2020-10.03.2020 (Figure 2). At sites B to E all MP tested SARS-CoV-2 negative. Of note, site B provided another 4 MP artificially spiked with SARS-CoV-2 positive NA samples from actual cases to further validate the procedure. The Ct-values of SARS-CoV-2 RT-PCR in individual patient samples were 26, 26, 15, and 35, respectively. All artificially spiked MP tested SARS-CoV-2 positive and Ct-values were 29, 29, 18, and 38 indicating a dilution factor of 10 as expected.

**Table 2:**
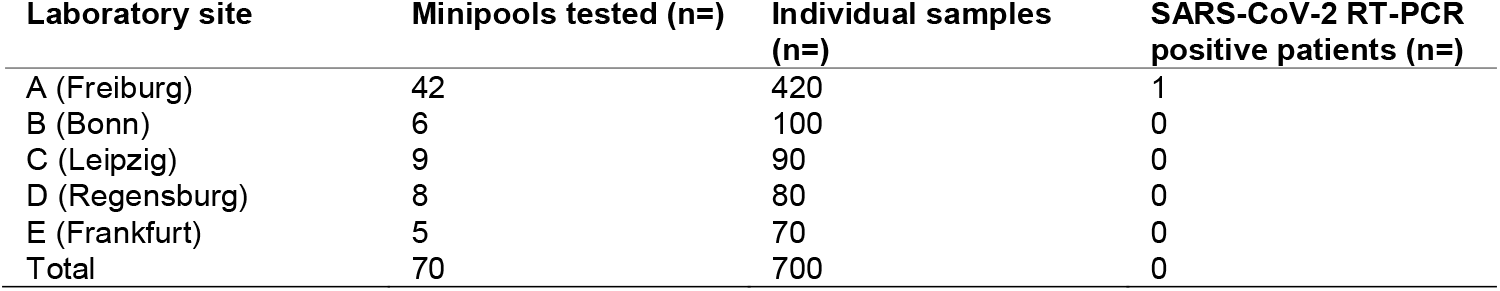
Number of minipools tested for SARS-CoV-2 RNA at five different sites, Germany, February – March 2020 (n=60).

## Conclusions

We report a diagnostic workflow for the laboratory-based surveillance of SARS-CoV-2 in a rapid and cost effective manner. Shortly after the identification of SARS-CoV-2 specific real-time RT-PCR protocols were set up and have been distributed worldwide (4, 5). The availability of rapid and reliable diagnostics for early case detection is instrumental in an outbreak scenario (6). From a public health perspective an easy to establish and cost effective laboratory-based screening strategy may assist in rapid case detection and ultimately in a better understanding of this epidemic (7). Technically, this can be done in parallel using samples from routine diagnostics which are subsequently tested for SARS-CoV-2 RNA (8). However, with the circulation of influenza cases across Europe merging with the upsurge of SARS-CoV-2 many laboratories may lack the capacity and resources to perform additional single patient sample testing for SARS-CoV-2. In addition, a shortage of PCR reagents has become an issue of concern as huge numbers of additional SARS-CoV-2 molecular tests are performed globally in a relatively short period of time. To minimize work load, resources and costs a pooling approach of nucleic acid extractions might be considered. We used the assay described by Corman et al. and were able to demonstrate an almost exactly 10-fold higher LOD which is due to MP related dilution factor of 10 (4). Data from China showed SARS-CoV-2 RNA concentrations in the range of 1,5×10^4^ to 1,5×10^7^ copies per milliliter giving rise to the notion that the MP procedure will be sensitive enough for most clinical samples (9). However, at the moment there is a lack of comprehensive information on viral RNA concentrations in mildly diseased or asymptomatic cases. Critically, we were not able to detect one low concentrated samples diluted into a MP, which was close to the LOD of the pooling procedure.

Networks are paramount for an efficient response to emerging infections and we aimed to provide an easy to implement workflow (4, 10). We set up an informal network and were able to test a total of 70 MP covering different geographic regions of Germany. In perspective, this approach can be set up rather easily e. g. by public health laboratories, can be done on a daily basis and at reduced costs compared to individual patient testing. It could allow for longitudinally monitoring the effectiveness of contact reduction measures at the population level and early detection of epidemic waves.

In light of an evolving SARS-CoV-2 epidemic and the possibility of unrecognized spread within the population we propose a rapid and straightforward screening strategy for SARS-CoV-2. This approach proved its principle and might assist public health laboratories in Europe and elsewhere to rapidly detect SARS-CoV-2 cases which might otherwise remain undetected.

## Data Availability

The data that support the findings of this study are available from the corresponding
author upon reasonable request.

## Acknowledgement

We are grateful to Claudia Ehret, Monika Häffner, Verena Schillinger and the team in Freiburg and the entire molecular diagnostic teams in Bonn, Frankfurt, Leipzig, and Regensburg for expert technical assistance.

## Ethical considerations

All samples have been submitted for routine patient care and diagnostics. Ethical approval for this study was not required since all activities are according to legal provisions defined by the German Infection Protection Act (IfSG). Written informed consent has been obtained by each patient. All data used in the current study was anonymized prior to being obtained by the authors.

## Data availability

The data that support the findings of this study are available from the corresponding author upon reasonable request.

